# Pathways of the COVID-19 Pandemic with Human Mobility across Countries

**DOI:** 10.1101/2020.05.21.20108589

**Authors:** Cheng Zhang, Li-Xian Qian, Jian-Qiang Hu

## Abstract

This study develops a holistic view of the novel coronavirus (COVID-19) transmission worldwide through a spatial-temporal model with network dynamics. By using a unique human mobility dataset containing 547,166 flights with a total capacity of 101,455,913 passengers among 22 countries during the past three months, we analyze the associations between international travel movement in six continents and the new infections in these countries. Results show that policymakers should move away from the previous practices that focus only on restricting hotspot areas with high transmission rates. Instead, they should develop a new holistic view of global human mobility to adjust the international movement restriction. The study highlights that international human mobility is the key to understand the transmission pathways and the small world phenomenon in the global network of COVID-19 pandemic.

## Introduction

Although countries worldwide have gradually restricted non-essential international travel across countries, particularly those from high-transmission areas, it does not seem to have had the desired effect of stopping the spread of the novel coronavirus (COVID-19) in the past few weeks. The epidemic continues to be severe in the European and American regions. It has even gradually spread to more areas, such as Africa, leading to controversy over the pathway of the epidemic transmission ^1,2^ The urgent situation needs a better assessment of the spread of the COVID-19 under global human mobility ^3,4^

Besides non-pharmaceutical interventions within each country ^5^, restrictions on non-essential international travel from epidemic areas are proposed as a critical strategy to slow transmission^6^. It intends to cut off people’s outbound movement from a country when it becomes a hot spot of the epidemic. This approach may not be as effective as policy makers expect if they ignore the current reality of global human mobility. Another important decision-making factor that has been overlooked is how to impose internal movement control policies, such as curfew or other forms of travel restriction, in the infected areas. These issues are particularly important in designing a global strategy to respond to the COVID-19 dynamics and to recover social and economic activities.

Figure 1 below summarizes three different views on understanding the role of human mobility in the pandemic. The first is a focal view that addresses the role of internal mobility and movement controls in tackling epidemic in a single country while ignoring the influence of inbound and outbound mobility ^7-10^. The second is a dyadic view that explores the role of international mobility and travel restriction between a centric country (usually a hotspot area in the epidemic) and some other countries ^11-13^. This approach usually does not take into account the impact of population movements other than those of the centric country, and in addition, only the one-way impact of the centric country on other countries. The third is a holistic view that analyzes the multiple paths of international mobility and travel restrictions in a global network simultaneously and allow for the correlation of each path in the network. In this way, the networked approach allows the study of simultaneous and asymmetric effects between multiple countries through different mobility pathways.

**Figure 1.**
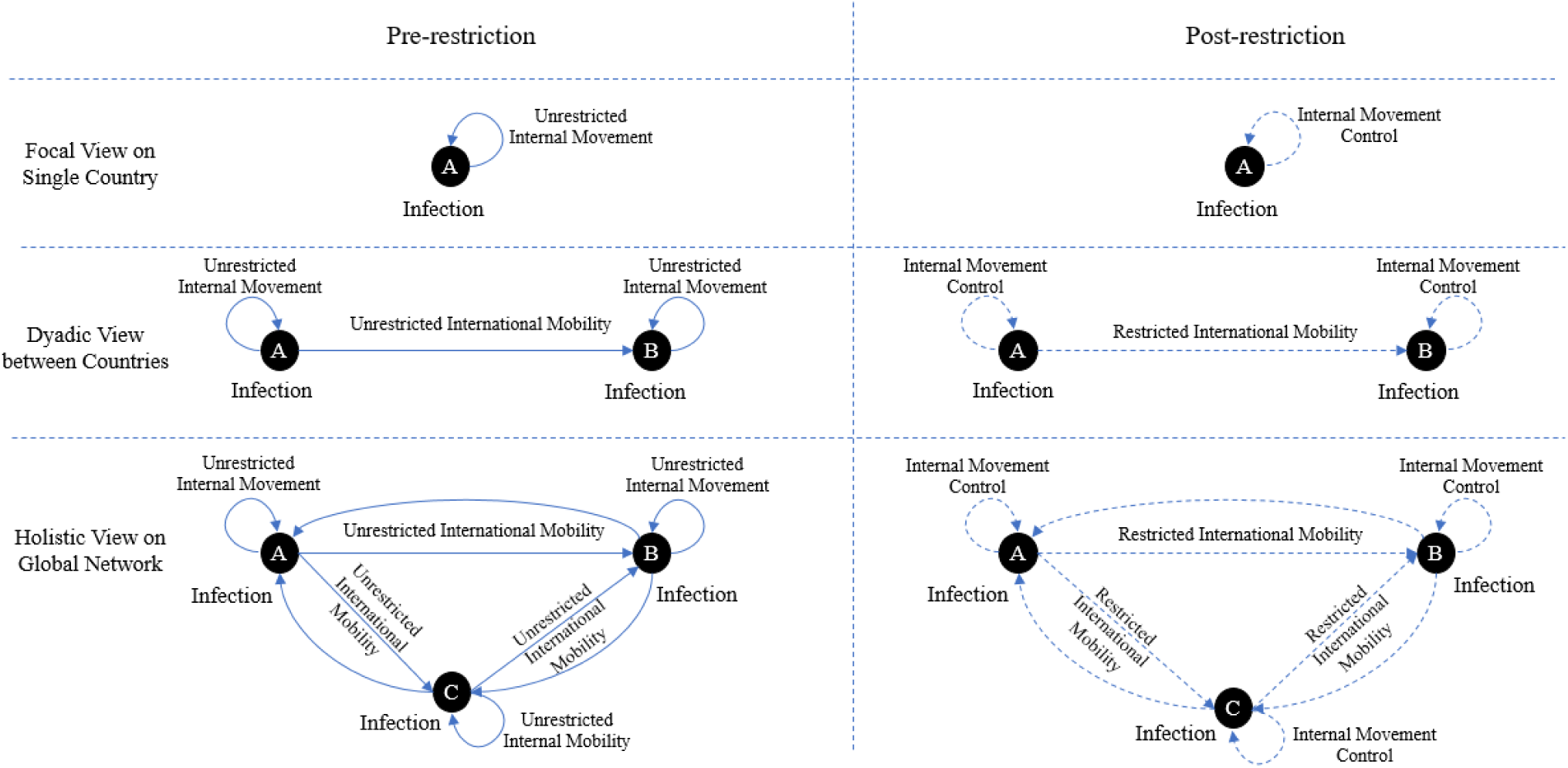
Three views on the role of human mobility in the pandemic. Note: The solid and dashed lines indicate unrestricted human mobility before public interventions and restricted mobility after public interventions, respectively. A line with the arrow from country A to country B stands for the epidemic influence from A to B due to (restricted or unrestricted) international mobility. An arrow connector on a country itself represents the internal transmission of epidemic due to (controlled or unrestricted) internal movement.

Global network-based analysis is now more urgently needed than previous focal view approach^7-10^ or dyadic view approach^11-13^. That is, investigating transmissions between countries in the global network where each country is a node of the network and the intercountry human mobility represents the edge of the network. In this way, scholars and policy-makers can understand the holistic pathways of the virus’ spread across the globe and develop effective worldwide-coordinated measures for the pandemic^14^.

This study responds to this urgent calling by developing a spatial-temporal model with network dynamics ^15,16^ to understand the inter-country transmission of the COVID-19 between January 22, 2020, when the epidemic was officially reported by the World Health Organization (WHO), to April 24, 2020. Based on the timing of the outbreak and the scale in terms of the accumulated confirmed infection numbers in the COVID-19 pandemic, we selected 22 countries from six continents, which accounted for 86.70% of total infection amount worldwide as of April 24, 2020. Detailed information of the countries is provided by Table S1 in supplementary materials.

We use the daily number of confirmed COVID-19 infections in every country from the WHO^1^. We also use a unique dataset containing 547,166 flights with a total capacity of 101,455,913 passengers among the 22 countries during the past three months. In the follow-up analyses, the information is aggregated to the country-day level, resulting in 23,885 daily pairs of international travel movement across six continents. Figure S1 illustrates a clear decreasing trend since early March regarding the capacity and number of international flights among these 22 countries. We also compile data from multiple sources containing international travel restriction and internal movement control policies (such as curfew and other forms of domestic travel restriction), as summarized in Table S1.

By treating each country as a node in the global network and the international human mobility as the connections between countries, we model the dynamic process of the epidemic. The unit of analysis of the study is the number of confirmed infections in country *i* on day *t*. At the beginning of the epidemic, every country has zero infection and human mobility is unrestricted between countries. Likewise, there is neither non-pharmaceutical interventions within countries nor international mobility restriction among countries. Therefore, countries around the world have formed a fully connected human mobility network.

During the pandemic period, however, travel restrictions were introduced by countries gradually and applied to travelers from different countries at different times, making the global human mobility network dynamically changing. The restriction reduces the human mobility between countries. If the action, saying the restriction from a hotspot country A to country B, takes effect, we would expect to see a weaker association between the epidemic in country A and the daily new cases in country B in the model compared to that in the period before the restriction. This would effectively imply a decreasing epidemic transmission from country A to country B, largely due to the fewer number of travelers from country A to country B.

However, reducing the spread of the epidemic does not solely depend on the international travel restriction, but also relies on the effective internal movement control within each country. Specifically, even if country B reduces international movement from the hotspot country A, the virus may still have chance to spread within country B (e.g., via internal transmission or from the third country), so that the impact of the epidemic in country B cannot be reduced without effective internal movement control policies. Therefore, if country B effectively implements internal movement control, we would also expect the control to work with international mobility restriction and observe a decreasing epidemic trend in country B, after a certain lagged period.

This idea is applied to all the countries in the network, taking into account the multiple paths of international mobility and travel restrictions between any two of them simultaneously. Through such a global network perspective, we aim to examine the extent to which the human mobility and the introduction of international travel restriction targeting hotspot countries correlate with the numbers of newly confirmed infections in other countries.

## Results

Table 1 presents the estimation results, where the red and blue colors highlight the positive and negative effects, respectively, both significant at 5% level. First, significant coefficients, no matter whether positive or negative, indicate clear associations of the epidemics between countries. Among them, the United States, South Africa, Switzerland, the Netherlands, Brazil are significantly affected by the outbreak in more than five countries worldwide. At the same time, the epidemics in the United States, Germany, United Kingdom, Turkey, and the Netherlands have significantly affected five or more other countries. Overall, these results indicate that the spread of the epidemic worldwide has revealed clear pathways over the past three months, where countries are influencing and influenced by different countries through various pathways in the network. Among them, the United States and the Netherlands show a clear hub effect of the transmission. The full estimation results are summarized in Table S2.

**Table 1:** Pathway Mapping of COVID-19 local and international transmission

|  | Dependent variable = daily new cases in each country |  |  |  |  |  |  |  |  |  |  |  |  |  |  |  |  |  |  |  |  |  |  |  |
| --- | --- | --- | --- | --- | --- | --- | --- | --- | --- | --- | --- | --- | --- | --- | --- | --- | --- | --- | --- | --- | --- | --- | --- | --- |
| Variable | USA | ESP | ITA | FRA | DEU | GBR | TUR | IRN | CHN | RUS | BRA | BEL | CAN | NLD | CHE | IND | PRT | ECU | JPN | KOR | AUS | ZAF |  |  |
| Intercept |  |  |  |  |  |  |  |  |  |  |  |  |  |  |  |  |  |  |  |  |  |  |  |  |
| 1-day lagged new cases ($\alpha_1$) | | | | | | | | | | | | | | | | | | | | | | | | |
| 14-day lagged pre-LocalControl ($\alpha_2$) | | | | | | | | | | | | | | | | | | | | | | | | |
| 14-day lagged international transmission of COVID-19 from the country in row head to the one in column head ($\alpha_3$) | | | | | | | | | | | | | | | | | | | | | | | Positive | Negative |
| CI_USA |  |  |  |  |  |  |  |  |  |  |  |  |  |  |  |  |  |  |  |  |  |  | 3 | 2 |
| CI_ESP |  |  |  |  |  |  |  |  |  |  |  |  |  |  |  |  |  |  |  |  |  |  | 0 | 2 |
| CI_ITA |  |  |  |  |  |  |  |  |  |  |  |  |  |  |  |  |  |  |  |  |  |  | 1 | 2 |
| CI_FRA |  |  |  |  |  |  |  |  |  |  |  |  |  |  |  |  |  |  |  |  |  |  | 0 | 4 |
| CI_DEU |  |  |  |  |  |  |  |  |  |  |  |  |  |  |  |  |  |  |  |  |  |  | 4 | 1 |
| CI_GBR |  |  |  |  |  |  |  |  |  |  |  |  |  |  |  |  |  |  |  |  |  |  | 5 | 1 |
| CI_TUR |  |  |  |  |  |  |  |  |  |  |  |  |  |  |  |  |  |  |  |  |  |  | 1 | 4 |
| CI_IRN |  |  |  |  |  |  |  |  |  |  |  |  |  |  |  |  |  |  |  |  |  |  | 0 | 0 |
| CI_CHN |  |  |  |  |  |  |  |  |  |  |  |  |  |  |  |  |  |  |  |  |  |  | 0 | 0 |
| CI_RUS |  |  |  |  |  |  |  |  |  |  |  |  |  |  |  |  |  |  |  |  |  |  | 0 | 1 |
| CI_BRA |  |  |  |  |  |  |  |  |  |  |  |  |  |  |  |  |  |  |  |  |  |  | 1 | 2 |
| CI_BEL |  |  |  |  |  |  |  |  |  |  |  |  |  |  |  |  |  |  |  |  |  |  | 1 | 1 |
| CI_CAN |  |  |  |  |  |  |  |  |  |  |  |  |  |  |  |  |  |  |  |  |  |  | 2 | 1 |
| CI_NLD |  |  |  |  |  |  |  |  |  |  |  |  |  |  |  |  |  |  |  |  |  |  | 1 | 4 |
| CI_CHE |  |  |  |  |  |  |  |  |  |  |  |  |  |  |  |  |  |  |  |  |  |  | 2 | 1 |
| CI_IND |  |  |  |  |  |  |  |  |  |  |  |  |  |  |  |  |  |  |  |  |  |  | 1 | 1 |
| CI_PRT |  |  |  |  |  |  |  |  |  |  |  |  |  |  |  |  |  |  |  |  |  |  | 2 | 1 |
| CI_ECU |  |  |  |  |  |  |  |  |  |  |  |  |  |  |  |  |  |  |  |  |  |  | 1 | 0 |
| CI_JPN |  |  |  |  |  |  |  |  |  |  |  |  |  |  |  |  |  |  |  |  |  |  | 0 | 0 |
| CI_KOR |  |  |  |  |  |  |  |  |  |  |  |  |  |  |  |  |  |  |  |  |  |  | 0 | 0 |
| CI_AUS |  |  |  |  |  |  |  |  |  |  |  |  |  |  |  |  |  |  |  |  |  |  | 1 | 1 |
| CI_ZAF |  |  |  |  |  |  |  |  |  |  |  |  |  |  |  |  |  |  |  |  |  |  | 4 | 0 |
| System Weighted R-Square | 0.9314 |  |  |  |  |  |  |  |  |  |  |  |  |  |  |  |  |  |  |  |  |  |  |  |
| Direct routes to the country in column head | 20 | 19 | 18 | 20 | 19 | 21 | 19 | 10 | 19 | 16 | 13 | 14 | 18 | 20 | 17 | 16 | 14 | 6 | 16 | 14 | 8 | 11 |  |  |
| Count of positive associations | 3 | 1 | 1 | 2 | 3 | 1 | 0 | 0 | 1 | 0 | 3 | 1 | 1 | 3 | 4 | 0 | 1 | 0 | 0 | 0 | 1 | 4 | 30 |  |
| Count of negative associations | 7 | 1 | 0 | 2 | 1 | 1 | 1 | 0 | 0 | 0 | 3 | 1 | 1 | 3 | 3 | 0 | 0 | 0 | 2 | 0 | 0 | 3 |  | 29 |

Positive effect ( $p < 0.05$ )
Negative effect ( $p < 0.05$ )
No direct international flights from the country in row head to the one in column head
Note: the abbreviations for country names are provided in Table S1

Following the typical setting in a network model, where value 1 is commonly used to indicate a connection between two nodes and 0 indicates no connection, we introduce a dummy variable with value 0 for the period with the international mobility restriction and value 1 to indicate the regular travel or connection between the two countries is allowed. Likewise, another dummy variable is used to set the internal movement control period as 0 and the pre-control period as 1 when the internal movement is fully allowed in the country. Given these settings, the positive coefficients in the estimation outcome show that the international transmission from country A to B in the pre-controlled period is stronger than in the post-restriction period. In other words, the positive coefficients show the effectiveness of the policies on reducing international transmission. Conversely, the negative coefficients show that the international transmission from country A to B in the pre-restriction period is weaker than in the post-restriction period. That means, the international transmission becomes stronger even after introducing the international travel restriction policies. Similarly, for internal movement control policies, the positive coefficients mean the more severe epidemic in the pre-controlled period than in the controlled period, which thus implies the effectiveness of internal movement control. On contrary, the negative coefficients would suggest the ineffectiveness of internal movement control policy in the corresponding country. Table S3 provides the implications of positive and negative effects of these two types of restriction interventions.

As shown in Table 1, the epidemic in China is largely isolated from those of the remaining world. That is, the number of new infections in China is not significantly associated with the epidemics in other countries except Italy (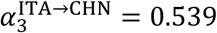, *p* < 0.001). The epidemic in China does not show any strong associations with the outbreaks in the rest 21 countries either. That is, all the associations between China’s cumulative infection number and the outbreak in other 19 countries with direct flights are insignificant (Note that China has no direct flights to Brazil and Ecuador). The main reason may be due to China’s early restriction on internal movement and its restriction on non-essential international travel outbound from China in late January. Travel restrictions imposed on China by many other countries in February and March might also help control international transmission from China. Other similar cases are South Korea and Iran, in which the epidemics have been well isolated, as their infection numbers are not associated with other countries’ lagged epidemics. The difference between Iran and South Korea/China is that there are fewer international direct flight routes to/from Iran, which might reduce the international transmission related to Iran.

However, other Asian countries such as Japan and India experienced a different situation as they have stronger connections with other countries. The epidemic in Japan is not significantly associated with other countries’ outbreaks but is associated with the lagged epidemic outbreaks from United States (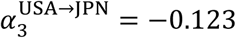, *p* < 0.05) and Russia (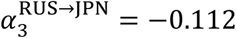, *p* < 0.05). Although the outbreak of COVID-19 in India was quite late, its epidemic has a significant association with the outbreaks in European countries, such as Italy (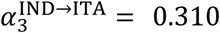, *p* < 0.05) and the Netherlands (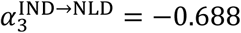, *p* < 0.01).

The results also show that the outbreak in the United States is largely associated with countries in Europe, South America, and Africa, rather than Asia. Specifically, we find that the cumulative infections in Germany, Portugal, and South Africa are positively associated with the new infections in the United States (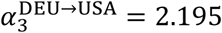, *p* < 0.001; 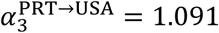 *p* < 0.01; 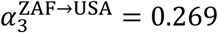, *p* < 0.01), which suggest the transmissions from these countries to the United States have been well controlled after introducing international travel restrictions in March. In comparison, even with the restricted international travels to the United States, its new infections are found to have stronger association with more countries than in the pre-restriction period, which are Spain, Italy, France, the United Kingdom (UK), Belgium, Brazil, and Turkey (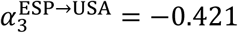, *p* < 0.001;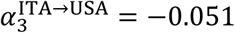, *p* < 0.05; 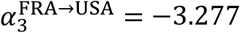, *p* < 0.001;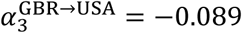, *p* < 0.05; 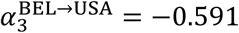, *p* < 0.05;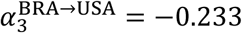, *p* < 0.05; 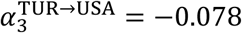, *p* < 0.01). Besides, the epidemic in the United States is associated with the outbreak in five countries, which are Germany, UK, Canada, South Africa, and Japan. As another North American country, Canada is found to have strong associations with the epidemics in France, Australia, the Netherlands, Turkey, in addition to the United States. Overall, the North American countries have the strongest association with European countries, followed by those in Africa, South America, and Asia.

We further find a close circle of mutual transmissions among European countries. Taking Spain as an example, its outbreak is found to have been strongly associated with those in other European countries, such as the UK (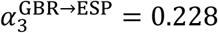, *p* < 0.001) in early-stage, followed by Italy (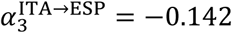, *p* < 0.05) more recently. What’s more, the epidemic in Spain positively associates with outbreaks in the Netherlands and the United States. Similarly, the outbreak in Germany is significantly associated with the epidemics in the UK and the Netherlands in Europe as well as the United States and South Africa outside Europe. Meanwhile, the epidemic in Germany also has strong associations with the transmission to Belgium, Switzerland, the United States, Brazil, and South Africa.

Our analysis also reveals the third wave epidemics differ in West Asia (e.g., Turkey), East Europe (e.g., Russia), South America (e.g., Brazil and Ecuador), and Africa (e.g., South Africa). Specifically, the outbreak in Turkey is associated with the epidemic in Canada (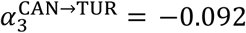, *p* < 0.01), and its epidemic is further associated with the transmission to more countries, such as France, Switzerland, United States, Brazil and South Africa. The epidemic in Russia is significantly associated with the recent upsurge of the COVID-19 epidemic in Japan (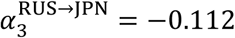, *p* < 0.05). In comparison, the outbreak in South America is strongly associated with Germany, UK, the Netherlands and France in Europe as well as the United States, given their close connections with direct flights. In addition, South Africa has a strong bilateral association with the United States on the transmission of COVID-19, and its outbreak is strongly influenced by the epidemics in Europe. At the same time, the epidemic in South Africa is also associated with the outbreak in Australia.

In summary, our analysis provides insightful findings on the correlations of the COVID-19 pandemic across countries. Figure 2 illustrates the global transmission map based on our estimation results. The size of each node represents the confirmed infection amount in the corresponding country by April 24, 2020. A red (blue) line represents the positive (negative) association between international movement and countries’ new infections with the strength of the line representing the value of the standardized correlation coefficient and an arrow showing the direction. Figure 3 further illustrates the overall network pattern of the virus transmission across different countries in different continents (as designated in different colors).

**Figure 2:**
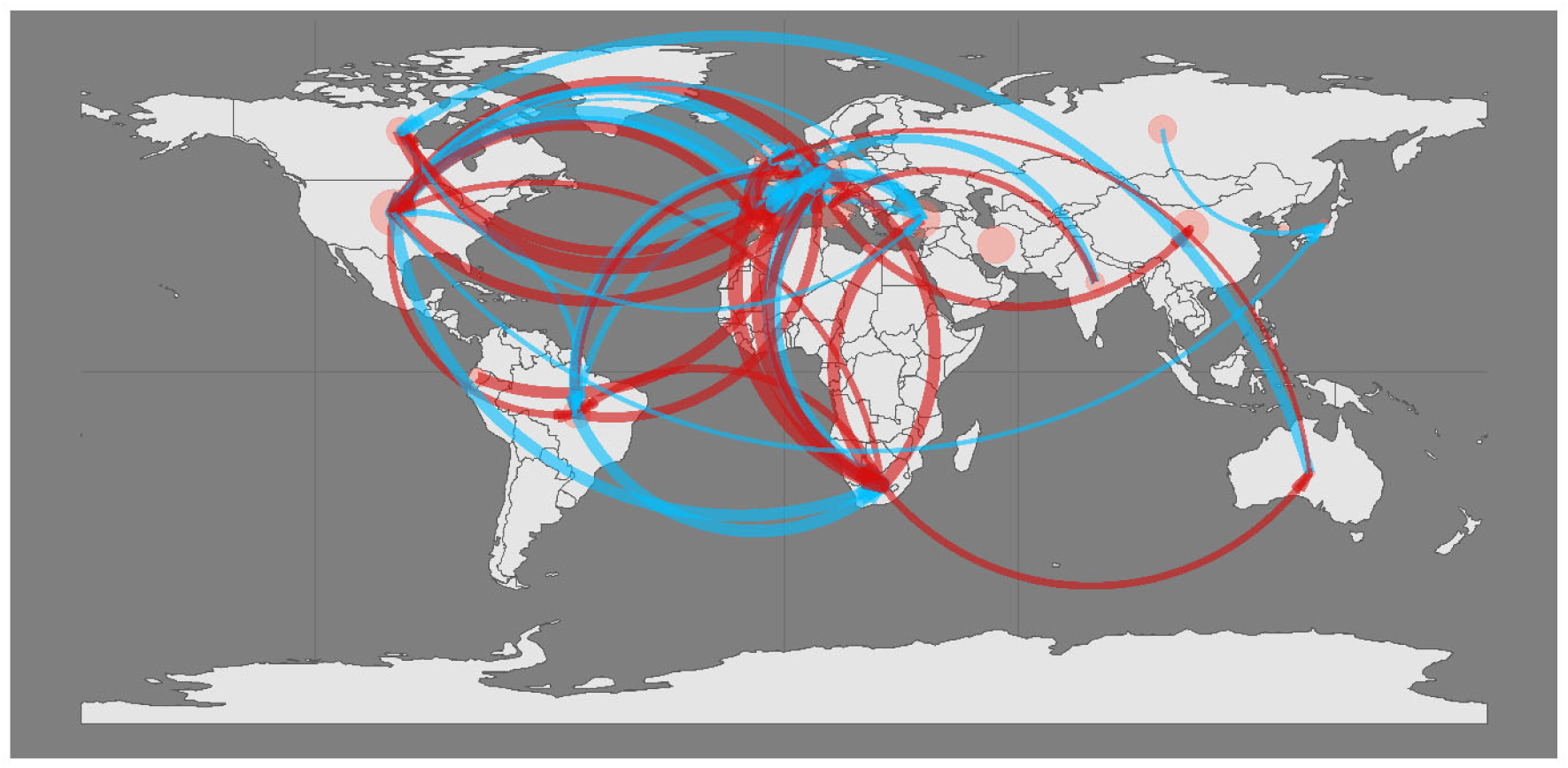
Transmission map of the COVID-19 pandemic among 22 countries. Note: the size of bubble of each country represent infection amount.

**Figure 3:**
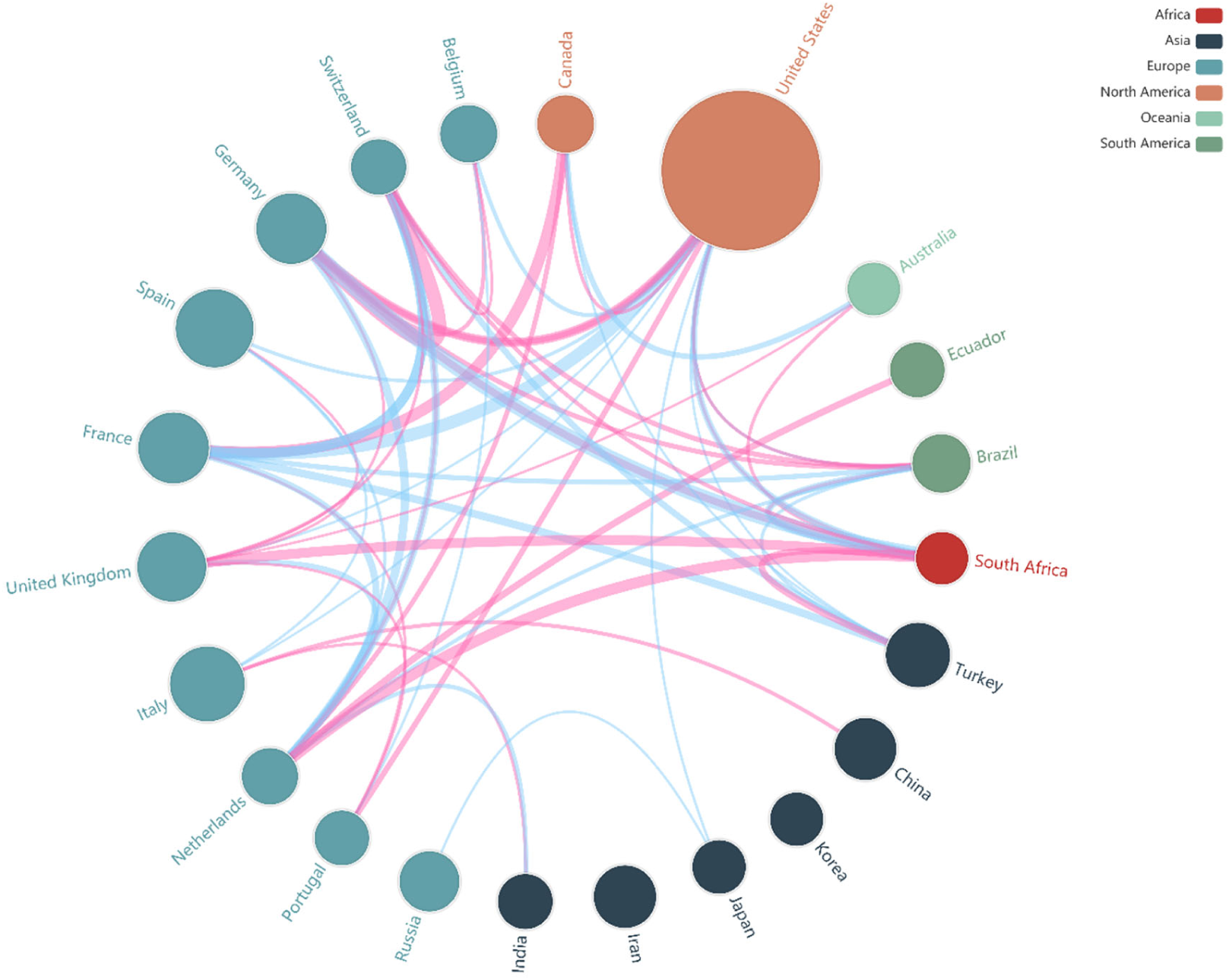
Continent-country level transmission.

Furthermore, based on the estimation results of the spatial-temporal model with network dynamics, we calculate the network properties of the countries, including betweenness centrality, closeness centrality, Eigenvector centrality and clustering coefficients^17–19^, where the countries are the node and the pathways are the edges between the nodes in the network. Table S4 presents the calculation results. Betweenness centrality of a node is the number of times a node acts as a bridge along the shortest path between two other nodes in a network. A high betweenness of a node reflects its high probability to occur on a shortest path between any two nodes. The findings imply that the United States, the Netherlands and Italy act as such top 3 bridges to facilitate the international transmission of the pandemic in the global network over the past three months. The closeness centrality of a node is the average length of the shortest path between the node and all other nodes in a network. A high closeness of a node reflects its closeness to other nodes in a network. The findings suggest that the United States, the Netherlands, United Kingdom and South Africa are such countries that closely link to the rest countries in the international transmission of the pandemic. Similarly, Eigenvector centrality of a node measures its relative importance in term of the influence to other nodes in a network, based on the concept that connections to high-importance nodes contribute more to the importance of the node. A high eigenvector score means that a node is connected to many nodes who themselves also have high eigenvector scores. In Table S4, the United States, the Netherlands and South Africa are the top-3 influential countries in the international transmission of the pandemic. All indicators taken together, the United States and the Netherlands play a most crucial role in the international transmission network of the pandemic.

A clustering coefficient is a measure of the degree to which nodes in a network tend to cluster together rather than being randomly connected between them. In other words, the clustering coefficient quantifies how close a node and its neighbors are to be a clique. Table S4 suggests that European countries including France, Switzerland, Germany and Belgium and the countries in the third-wave including South Africa, Brazil and Turkey are more likely to be such cliques with their neighbors, given their larger clustering coefficients. Another role of the clustering coefficient is to measure whether a network is a random or a small world ^19^ Following the equation that C_*random*_ = k/*n*^2^ given the n nodes and k total connections in between, we can get the clustering coefficient C_*random*_ = 0.122 in a random global network with 22 countries as nodes and 59 connections as edges. The value is smaller than the average clustering coefficient, i.e. 0.211, in the actual transmission network, suggesting that the international transmission network tends to be a small-world. That is, a sparse and decentralized network that is neither completely ordered nor completely random but with the pattern of high local clustering^20^. We argue that the international human mobility is the key to such network formation.

To check the robustness of our results, we repeat our analysis by using the number of international flights instead of flight capacity in the model. The results are consistent with that in the main analysis and summarized in Table S5. We further consider the situation without flight information available by replacing the flight capacity amount with 1/0 to indicate whether a country implemented international travel restriction with another country or not on a specific day. The results in Table S6 show that the patterns and pathways we observe are largely held unchanged and may simulate the situation of international travels with transitions in third countries. Finally, we replicate the analysis by using another COVID-19 dataset maintained by the Center for Systems Science and Engineering (CSSE) at Johns Hopkins University ^21^ which contains more sources of information from CDC and online media sources in many countries. Results show largely consistent pattern of the pathways across countries and continents, although country-specific impact coefficients vary in the global network. For instance, China is still found to be fully independent from the epidemics in all other countries. Also, the United States is found to be consistently influenced by the European countries. The results based on the data from Johns Hopkins University are presented in Table S7 to S9, corresponding to three models with the international flight capacity, the number of international flights and without flight information.

Considering potential collinearities between the estimated coefficients, we conduct a step-wise approach that adds countries’ variables continent by continent. Using the United States as an example, the stepwise analysis in Table S10 shows the consistent effects of international transmission, that is, the impacts from other countries’ epidemics on the daily new infections in the United States remain largely same and robust regarding the effect sizes and significance levels.

Our analysis also shows that the 14-day internal movement restriction policy is insufficient in controlling the transmission of COVID-19, given that the coefficient (*α*_2_) is negative in 19 countries and even significantly negative in 17 ones. See the 17 blue cells in the row of “14-day lagged pre-LocalControl” in Table 1. This coefficient is positive and insignificant for China, South Korea, and Australia. Further sensitivity analysis shows that this conclusion does not change much with 21 days’ lag for internal movement restriction. However, the introduction of 28 days’ lag would make three countries to have a positive and significant effect on this policy, which are China, Iran, and Switzerland, with positive despite the insignificant effect on more countries, including France, Germany, Turkey, the Netherlands, South Korea, and Australia. See Table S11 for the effects of the 21 days and 28 days of internal movement control policy.

## Discussions

By using the global human mobility data, this study provides the first comprehensive insights into the characteristics of international transmission in the COVID-19 pandemic. First, the study reveals clear pathways of international transmission among countries over the past three months. Second, the study identifies the hub effect of the United States and the Netherlands in the pandemic. A comprehensive network analysis further confirms that the United States and the Netherlands play a crucial role in the international transmission network of the pandemic.

Although the epidemic started in Asia first, the study finds that Asian countries take diverse roles in the successive transmission. For example, China, which was the first country to face the epidemic and was severely affected, has almost isolated itself from the international transmission network. In contrast, other Asian countries, such as Japan, show significant connections with other countries. The study further reveals that the epidemic in North American countries has the strongest association with European countries (followed by those in Africa, South America, and Asia). At the same time, there exists a close clique of mutual transmissions among many European countries. The network analysis further confirms such cliques among European countries and the third-wave countries including South Africa, Brazil and Turkey.

The first important takeaway from the study is the importance of a holistic view of the COVID-19 transmission. Restrictions on non-essential international human mobility between countries are one critical strategy to fight against epidemic outbreaks ^6^; however, little was known before regarding the proper ways to implement the strategy. As such, policymakers might focus only on the hotspot areas with a high transmission rate (which is not wrong) but overlook the pathway effect beyond the epidemic areas in the global network. Unfortunately, over the past three months, we have observed such a narrow perspective repeatedly occurring in many countries, focusing only on cutting off hotspot outbreak areas. New York Governor Andrew Cuomo recently start to question where the coronavirus that hit New York state came from: “……We closed the front door with the China travel ban, which was right……but we left the back door open because the virus had left China by the time we did the China travel ban”^22^ Through this study, we show that the establishment of a new holistic and networked view of pandemic transmission on a global scale is imperative, with the following important implications.

First of all, it cannot be simply assumed that the international travel ban from high-risk areas will be sufficient to control the epidemic once and for all while ignoring the dynamic spread of the epidemic in the global network. Policymakers in every country should keep abreast of population movements between countries and, through scientifically rigorous analysis, and foresee the pattern of epidemic transmission in the network. Accordingly, they should dynamically adjust the corresponding international restriction strategy promptly.

Second, the global perspective should also take into account the domestic movement restriction already implemented and its effects. Despite the early outbreak in Asian countries, their impact on other countries is not the same. Because of strict national and international restrictions imposed by China, the epidemic in China was developing in isolation: it didn’t affect other countries or was affected by other countries’ epidemics. In contrast, the lack of strict and timely domestic restriction in some other countries, combined with their lack of global view on timely international restriction, contributed to the pandemic, which means these countries were subsequently affected by the epidemic in the second and third waves.

By the same token, as epidemics are gradually brought under control in the coming months, the effects and extent of national and international movement restriction should both be taken into account when countries decide to resume international economic and social activities. At the same time, it’s also important to remain vigilant about the third-wave outbreak that may appear in Latin America and Africa. The holistic view should be applied promptly to guide the internal and international movement restriction policies in the areas.

Finally, our analysis also shows that there may exist factors outside the epidemic that are influencing countries’ decisions on international travel restrictions. For instance, when the United States started to ban non-US citizens or permanent residents who had been in China in the past 14 days to enter the United States on February 2, 2020, there were 16,640 infected patients in China with the infection rate was 11.95 per million people. In comparison, when the United States banned the entry of European people on March 13, Italy itself had 17,660 patients with an infection rate of 292.29 per million people. Therefore, this also adds another dimension of restriction timing that future research should be aware of.

As one of the first few studies to focus on a holistic view of the epidemic, the paper cannot avoid its limitations. First, all the restrictions were still in effect at the time of writing; therefore, we mainly focused on the short-term effects rather than a longitudinal examination. We recommend that scholars exercise caution when extrapolating our conclusions to longer periods. Second, some reliability concerns still exist related to the confirmed number reported and policy implementation; however, we could analyze only the available data. Furthermore, examining the restriction implementation process and efforts of the actions in a more detailed manner would increase the rigor and power of the initial analysis presented in this paper. Models that can predict the dynamic future is also the focus of the next steps.

## Methods

### • Data

We collected multi-sourced datasets in this study. The daily number of confirmed infections was collected from the WHO COVID-19 Dashboard. Based on the timing of the outbreak occurred and the scale in terms of their accumulated confirmed infection numbers during the outbreak, we selected 22 countries from six continents: 5 countries from Asia (China, Iran, India, Japan, and South Korea), 11 countries from Europa (Belgium, France, Germany, Italy, Netherlands, Portugal, Russian Federation, United Kingdom, Spain, Switzerland, and Turkey), 2 countries from North America (Canada, and United States), 2 countries from South America (Brazil, Ecuador), 1 country from Africa (South Africa), and 1 country from Oceania (Australia). These 22 countries accounted for 86.70% of the total infection amount worldwide as of April. 24, 2020. In addition to WHO data, we also collected data from and the COVID-19 Data Repository by the Center for Systems Science and Engineering (CSSE) at Johns Hopkins University ^21^ to replicate the analysis.

We further use a unique human mobility dataset containing daily global commercial flights among the selected 22 countries between January 22, 2020, and April 24, 2020, from a leading data consulting company in the civil aviation industry, VariFlight.^2^ The dataset contains information on the origin country, destination country, date, the number of flights, and total capacity in terms of the maximum available seats. In summary, this dataset provides information on 547,166 flights between 23,855 pairs of origin and destination countries that covers a total capacity of 101,455,913 passengers during the period.

We collected data on the international travel restriction between countries and the internal movement control within every country from Oxford COVID-19 Government Response Tracker^3^ and GardaWorld Crisis24 Global Portal^4^, which timely document epidemic prevention policies in countries around the world. The path connection from country A to country B is set by value 1 at day 0, representing the availability of international movement from A to B. Such connection is disabled on day *t* when either the internal movement control in country A or the entry ban for travelers from country A into B. Thus, on day *t* onwards, the connection from A to B is set to be 0 to represent the disconnection. Table S1 summarizes the timing and scale of the outbreak, as well as travel restriction policies by the 22 countries.

### • Dynamic Spatial Network Model

The unit of analysis of the study is the number of confirmed infections in country *i* on day t. Given the spatial nature of this research, we develop a dynamic network model based on the spatial-temporal model ^15,16^ to examine the extent to which (1) the mobility between countries, (2) the dynamics of travel restriction policies between countries, and (3) non-pharmaceutical interventions within each country, correlates with the numbers of newly confirmed infections in other countries. Specifically, the number of newly confirmed infections in the country *i* (*i* = 1 to 22) on day *t* (*t* = 1 to 94, starting at January 22 till April 24), *NCI_i,t_*, is given by

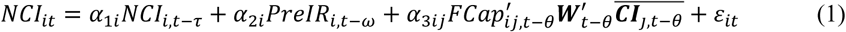

where the coefficient *α*_1_*_i_* measures the *τ*-day lagged effect of new infections in the same country, and *α*_2_*_i_* captures the *ω*-day lagged effect related to internal movement restriction, such as social distancing and home-stay orders, within country *i*, where

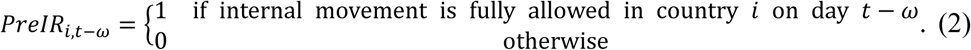

Specifically, the positive value of *α*_2_*_i_* represents the positive association between internal movement and the new infections in the country *i* after *ω* days. It also implies that the introduction of the movement restriction is negatively associated with the infection. 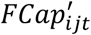 is the direct flight capacity, such as the number of direct flights or maximum flight seats in the direct flights from country *j* to *i* on date *t*, 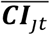 is the number of cumulative confirmed infections in each of the rest countries by day *t* (*j ≠ i*), and 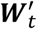 is the time-dependent spatial weight matrix of the international mobility network between all considered countries on day *t* with its element *w_ijt_* defined as

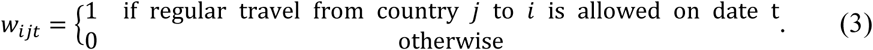

Specifically, the travel from country *j* to *i* on day *t* is conditional on both that the origin country *j* has not imposed the internal movement restriction on day *t* (so that regular travelers can leave country *j*) and that the destination country *i* has not banned the entry for travelers from country *j* (so that they can enter country *i* as well). Restriction in either the origin or the destination country would make the regular traveling difficult or even impossible. Therefore, *α*_3_*_ij_* measures the effect of international mobility between countries on the epidemics of COVID-19 in different countries by taking into account the *θ*-day lag that largely attributed to the incubation period. If *α*_3_*_ij_* >0, it represents the positive association between international transmission from country *j* to *i* and the newly confirmed infection in the country *i* after *d-*days. It also implies that the international travel restriction policy for the route from country *j* to *i*, is negatively associates with the newly confirmed infections in the country *i* after *θ*-days of its introduction. *ε_it_* is the error term.

### • Estimating the model

The econometric model we develop above has the time-dependent spatial weight matrix, 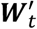. This unique feature makes our model differs from the conventional spatial models, including the dynamic spatial panel model, which typically use the time-invariant spatial weight matrix based on the geographical characteristics ^15^. Therefore, we follow the process below to estimate the model.

1. Code the elements of spatial weight matrix (*w_ijt_*), which may vary by the origin, destination, and date, based on the international travel information and internal movement control policies, as summarized in Table S1.
2. Organize the data into a time series format, with each row representing the date and each column for the different data variables on that date, such as the coded elements of the spatial weight matrix, the number of infection cases in every country and the numbers of flights and maximum available seats in every pair of international mobility route.
3. Create time-lagged variables where necessary. In the empirical analysis, we take *τ* = 1 to one-day lagged effect of new infections, *ω* = 14 to check the effect of internal movement restriction within each country, and *θ* = 14 to capture the influence of 14-day incubation period of the COVID-19 on its international transmission.
4. Construct an interdependent system of linear regression equations, and use Seemingly Unrelated Regression (SUR) ^23-25^ to estimate the parameters as specified in equation (1). The SUR is also known as joint generalized least squares (JGLS), and it is a generalization of OLS for multi-equation systems by allowing for the correlation of the error term of every equation. In an m-equation system for T-period observations,

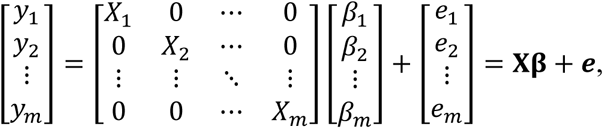 The variance-covariance matrix for the error term vector *e* can be written as

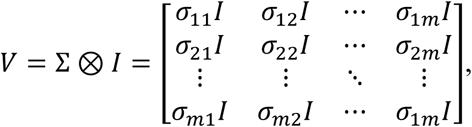

where ⊗ is the Kronecker multiplication operator, ∑ is an *m*\**m* positive definite symmetric matrix with *σ*_11_ as the variance of the error in the first equation and *σ_ij_* as the covariance between the errors of the *i^th^* and *j^th^* equations, and *I* is the identity matrix. The estimation process of SUR is as follows:
  a. First applies OLS to every equation and obtain the residual of every equation, *e_i_* (*i* = 1,2,…,m).
  b. Since Σ is typically unknown, the elements of Σ can then be estimated using

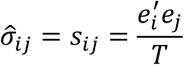
  c. Flexible Generalized Least Square (FGLS) estimators are used to estimate the coefficients.

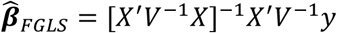

where [*X′V*^−1^*X*]^−1^ is the variance-covariance matrix of the estimated coefficients.

The model is implemented using the SUR estimator in the SYSLIN procedure in SAS. The SAS code is available in supplementary materials.

## Data Availability

Data will be available upon request.

## Acknowledgement

This research was partially supported by the National Natural Science Foundation of China (No. 91846302, 71720107003, 71973107).

## Footnotes

1 WHO COVID-19 Dashboard (https://covid19.who.int/)

2 http://www.variflight.com/en/

3 https://www.bsg.ox.ac.uk/research/publications/variation-government-responses-covid-19

4 https://www.garda.com/crisis24

## Notes

### Competing Interest Statement

The authors have declared no competing interest.

## References

1 Gonzalez-Reiche, A. S. et al. Introductions and early spread of SARS-CoV-2 in the New York City area. medRxiv, 2020.2004.2008.20056929, doi:10.1101/2020.04.08.20056929 (2020).

2 Forster, P., Forster, L., Renfrew, C. & Forster, M. Phylogenetic network analysis of SARS-CoV-2 genomes. Proceedings of the National Academy of Sciences, 202004999, doi:10.1073/pnas.2004999117 (2020).

3 West, R., Michie, S., Rubin, G. J. & Amlôt, R. Applying principles of behaviour change to reduce SARS-CoV-2 transmission. Nature Human Behaviour, doi:10.1038/s41562-020-0887-9 (2020).

4 Bavel, J. J. V. et al. Using social and behavioural science to support COVID-19 pandemic response. Nature Human Behaviour, doi:10.1038/s41562-020-0884-z (2020).

5 Wang, C. et al. Evolving Epidemiology and Impact of Non-pharmaceutical Interventions on the Outbreak of Coronavirus Disease 2019 in Wuhan, China. medRxiv, 2020.2003.2003.20030593, doi:10.1101/2020.03.03.20030593 (2020).

6 Paules, C. I., Marston, H. D. & Fauci, A. S. Coronavirus infections—more than just the common cold. JAMA (2020).

7 Guan, W.-j. et al. Clinical Characteristics of Coronavirus Disease 2019 in China. New England Journal of Medicine, doi:10.1056/NEJMoa2002032 (2020).

8 Huang, C. et al. Clinical features of patients infected with 2019 novel coronavirus in Wuhan, China. The Lancet (2020).

9 Jia, J. S. et al. Population flow drives spatio-temporal distribution of COVID-19 in China. Nature, doi:10.1038/s41586-020-2284-y (2020).

10 Giordano, G. et al. Modelling the COVID-19 epidemic and implementation of population-wide interventions in Italy. Nature Medicine, doi:10.1038/s41591-020-0883-7 (2020).

11 Joseph T Wu, K. L., Gabriel M Leung. Nowcasting and forecasting the potential domestic and international spread of the 2019-nCoV outbreak originating in Wuhan, China: a modelling study. The Lancet DOI:https://doi.org/10.1016/S0140-6736(20)30260-9 (2020).

12 Li, R. et al. Substantial undocumented infection facilitates the rapid dissemination of novel coronavirus (SARS-CoV2). Science, eabb3221, doi:10.1126/science.abb3221 (2020).

13 Kraemer, M. U. G. et al. The effect of human mobility and control measures on the COVID-19 epidemic in China. Science, eabb4218, doi:10.1126/science.abb4218 (2020).

14 EDITORIAL. COVID-19: too little, too late? The Lancet 395 (2020).

15 Elhorst, J. P. Spatial econometrics: from cross-sectional data to spatial panels. (Springer, 2014).

16 Belotti, F., Hughes, G. & Piano Mortari, A. Spatial panel data models using Stata. The Stata Journal 17, 139–180 (2017).

17 Freeman, L. C. Centrality in social networks conceptual clarification. Social networks 1, 215–239 (1978).

18 Newman, M. E. J. The structure and function of complex networks. SIAM review 45, 167–256 (2003).

19 Watts, D. J. & Strogatz, S. H. Collective dynamics of ‘small-world’ networks. Nature 393, 440–442, doi:10.1038/30918 (1998).

20 Watts, D. J. Networks, Dynamics, and the Small - World Phenomenon. American Journal of Sociology 105, 493–527, doi:10.1086/210318 (1999).

21 Dong, E., Du, H. & Gardner, L. An interactive web-based dashboard to track COVID-19 in real time. Lancet Infectious Diseases (2020).

22 Schwartz, I. (https://www.realclearpolitics.com/video/2020/04/24/cuomothecoronavirusthatcametonewyorkdidnotcomefromchinaitcamefromeurope.html, 2020).

23 Zellner, A. An Efficient Method of Estimating Seemingly Unrelated Regressions and Tests for Aggregation Bias. Journal of the American Statistical Association 57, 348–368 (1962).

24 Greene, W. H. Econometric Analysis. (Prentice Hall. New Jersey, 2003).

25 Ajmani, V. Applied econometrics using the SAS system. (John Wiley & Sons., 2011).

